# Sodium channel blockers are associated with reduced dementia risk in late-onset unexplained epilepsy

**DOI:** 10.64898/2026.05.20.26353714

**Authors:** Carolina Ferreira-Atuesta, Kai Michael Schubert, Xin You Tai, Daniela Noain, Daria Skwarzynska, Matthias T. Wyss, Simon Schreiner, Hans H. Jung, Trey Hedden, Johan Zelano, Tony Marson, Gregory Y. H. Lip, Gashirai K. Mbizvo, Marian Galovic

**Author notes:** authors share senior authorship (Gregory Y. H. Lip, Gashirai K. Mbizvo, Marian Galovic). **One Sentence Summary:** In a global target trial emulation of >75 million adults, sodium channel blocker antiseizure medications were associated with a 27% lower hazard of incident dementia in late-onset unexplained epilepsy compared with levetiracetam.

## Abstract

Late-onset unexplained epilepsy is a potential harbinger of dementia, likely driven by network hyperexcitability that facilitates amyloid-β release and tau propagation. Dampening this activity with antiseizure medications offers a potential disease-modifying strategy, yet whether specific agents differentially alter this neurodegenerative trajectory remains unknown. Here, we emulated a target trial using global real-world federated data on patients with late-onset unexplained epilepsy to compare dementia risk across antiseizure monotherapies. Using data from over 75 million adults aged 55 years or older, we found that sodium channel blockers were associated with a 27% lower hazard of incident all-cause dementia (hazard ratio = 0.73, 95% confidence interval 0.61–0.88) and 34% lower hazard of Alzheimer’s disease (hazard ratio = 0.66, 0.49–0.88), compared with levetiracetam/brivaracetam. While the class effect was protective, individual agents such as phenytoin, carbamazepine, and lamotrigine showed divergent safety and efficacy profiles. We replicated these findings in both a Down syndrome cohort and the external National Alzheimer’s Coordinating Center dataset. Our results suggest that targeting neuronal excitability with sodium channel blockers is associated with lower risk of dementia, prioritizing the repurposing of these agents for dementia prevention trials.

## INTRODUCTION

Late-onset unexplained epilepsy (LOUE) is increasingly recognized not merely as a comorbidity of Alzheimer’s disease, but as a potential harbinger and driver of dementia (1–4). Individuals with LOUE face a 2- to 3-fold increased risk of dementia compared with the general population, pointing to a shared pathophysiological interface (5, 6). Emerging evidence identifies aberrant neural network activity as the central mechanism linking these conditions (7). Mesiotemporal hyperexcitability, observed in both LOUE and prodromal Alzheimer’s disease, has been shown in animal models to drive activity-dependent amyloid-β release and facilitate tau propagation, the two pathophysiological hallmarks of Alzheimer’s disease (3, 7–11). Interstitial amyloid-β levels are dynamically regulated by synaptic activity, demonstrating that the inhibition of action potentials, specifically via sodium channel blockade, profoundly reduces extracellular amyloid-β concentrations (12, 13). Thus, seizures may catalyze a vicious cycle of excitability and protein accumulation, accelerating the transition to overt dementia.

This mechanistic coupling suggests that antiseizure medications (ASMs) might possess disease-modifying potential by dampening synaptic hyperactivity in prodromal dementia (14–18). Yet, clinical translation of this hypothesis remains limited. Previous trials have focused almost exclusively on the SV2A ligand levetiracetam and on later stages of the disease, yielding mixed results (4, 6, 8–10). Moreover, as ongoing studies continue to prioritize levetiracetam (4), the long-term neuroprotective potential of other common ASM classes remains largely unexplored. Here, we address this critical knowledge gap using a target trial emulation framework applied to a real-world international health dataset. We aimed to determine whether specific ASM monotherapies differentially modify the risk of incident dementia in patients with newly diagnosed LOUE. These findings may not only guide optimal ASM selection in clinical practice but also prioritize specific compounds for repurposing, offering a rapid, cost-effective roadmap for future basic and clinical research aimed at dementia prevention.

## RESULTS

### Participants and propensity-score matching

To emulate head-to-head comparisons of ASM monotherapy strategies in newly treated LOUE, we assembled new-user monotherapy cohorts within the TriNetX network, rigorously aligning time zero, eligibility criteria, and follow-up across treatment arms (table S1–S7). Among 75,516,264 adults aged 55 years or older, 94,250 met the eligibility criteria for LOUE and were stratified into five mechanistic ASM groups and six individual ASM cohorts (Fig. 1). Mean age was comparable across groups (73–75 years, range 56–90), with sex distribution varying by treatment strategy (table S8). Baseline comorbidity burden was largely similar, except for the gabapentin cohort, which showed a higher prevalence of comorbidities (table S8). Propensity-score matching effectively addressed these imbalances, yielding matched cohort sizes ranging from 1,906 to 11,682 patients per arm (Fig. 1, table S6). After matching, fewer than 1% of covariates showed residual imbalance (i.e., a standardized mean difference >0.1) (19) in either mechanism-group or single-ASM comparisons, indicating that most variables remained well balanced after adjustment, ensuring comparability (table S12–S32, fig. S4–S12).

**Figure 1.**
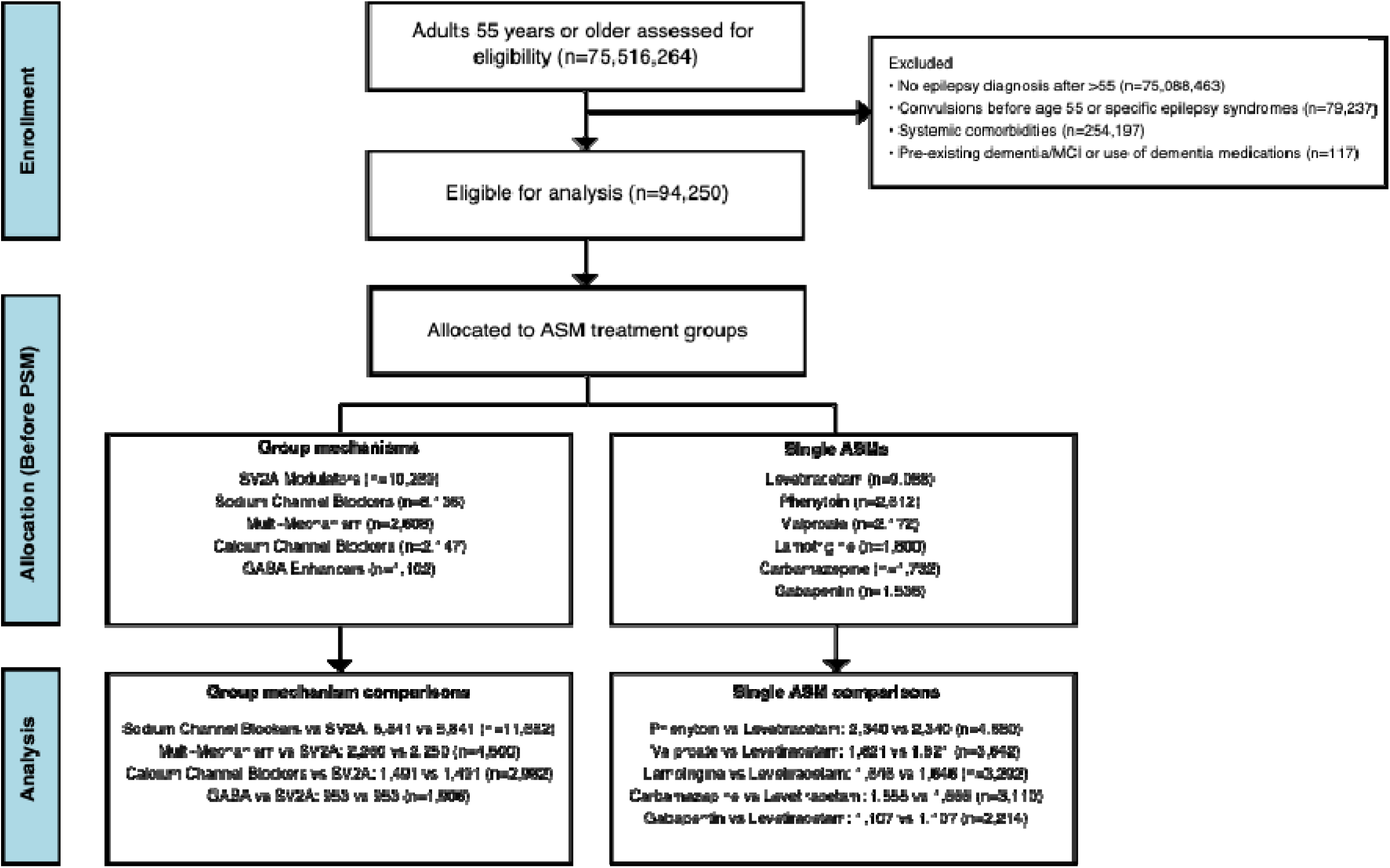
Flow Diagram. Patient flow from initial screening through propensity score matching. The diagram shows enrollment (initial assessment and exclusions), allocation to treatment groups, follow-up after 1:1 propensity score matching, and final analysis cohorts.

### Sodium channel blockers are associated with lower dementia risk in LOUE

We subsequently compared mechanistic ASM groups against synaptic vesicle glycoprotein 2A (SV2A) ligands levetiracetam or brivaracetam within matched cohorts to estimate the association between initial monotherapy selection and the risks of incident all-cause dementia, Alzheimer’s disease, and mortality. SV2A ligands were prespecified as the reference because levetiracetam is commonly prescribed and well-studied in LOUE, maximizing statistical power for active-comparator contrasts.

Sodium channel blockers were associated with a lower risk of incident dementia compared with SV2A ligands. Specifically, this class was associated with a 27% reduction in the hazard of all-cause dementia (hazard ratio [HR] = 0.73, 95% confidence interval [CI] 0.61–0.88, Fig. 2A) and a 34% reduction in the hazard of Alzheimer’s disease (HR = 0.66, 0.49–0.88, Fig. 2B). This corresponded to an estimated number needed to treat of 29 at 10 years for all-cause dementia and 48 for Alzheimer’s disease. Furthermore, we observed a 12% reduction in all-cause mortality (HR = 0.88, 0.78–1.00, Fig. 2C) and a 17% reduction in the composite endpoint of all-cause dementia or mortality (HR = 0.83, 0.74–0.94, Fig. 2D). Post hoc power analyses indicated adequate statistical power for the primary all-cause dementia comparison (90.1% power with 437 observed events; 325 events required for 80% power). For Alzheimer’s disease, the observed hazard ratio of 0.66 was supported by 185 events, yielding 80.5% post hoc power and exceeding the 183 events required to achieve 80% power.

**Figure 2.**
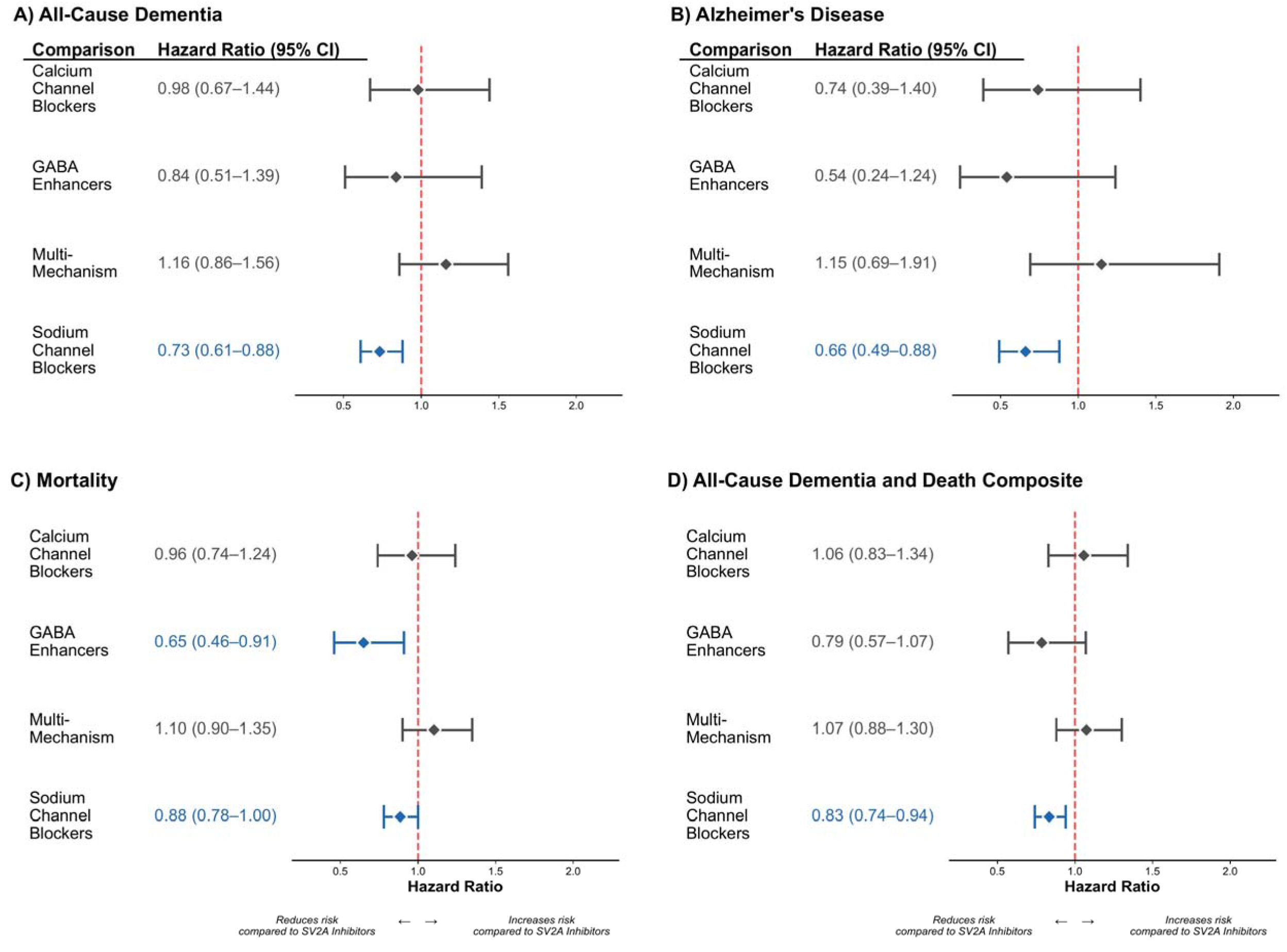
Forest plots of mechanism group comparisons on dementia risk. Hazard ratios (95% CI) for dementia and mortality outcomes comparing mechanism-based antiseizure medication groups versus SV2A inhibitors. Panels show: (A) All-cause dementia, (B) Alzheimer’s disease, (C) Mortality, (D) All-cause dementia + death. Diamond markers indicate hazard ratios; horizontal lines show 95% confidence intervals. Red dashed line at HR=1.0 indicates no difference. Blue indicates protective effect (CI upper bound <1.0), red indicates increased risk (CI lower bound >1.0), and gray indicates non-significant results. Per-protocol analysis with follow-up starting at day 366. Mechanism groups: Calcium channel blockers (Gabapentin, Pregabalin); GABA enhancers (Phenobarbital, Clonazepam, Clobazam); Multi-mechanism (Valproate, Topiramate, Zonisamide); Sodium channel blockers (Carbamazepine, Oxcarbazepine, Eslicarbazepine acetate, Lacosamide, Lamotrigine, Phenytoin); SV2A inhibitors (Levetiracetam). Secondary outcomes (MCI, vascular dementia, glaucoma) shown in supplementary figures.

Gamma-aminobutyric acid (GABA) enhancers were associated with a reduced risk of mortality (HR = 0.65, 0.46–0.91, Fig. 2C) but not with other outcomes, whereas calcium channel blockers and multi-mechanism agents showed no significant associations (Fig. 2).

### Replication in Down syndrome and the NACC cohort

To evaluate the generalizability of our primary findings to populations with distinct baseline risks and outcome ascertainment methods, we conducted (i) a validation in a TriNetX Down syndrome cohort and (ii) an external validation using the National Alzheimer’s Coordinating Center (NACC) dataset.

We identified 10,548 individuals with Down syndrome, creating mechanistic and individual-ASM cohorts with exposure definitions identical to the LOUE analysis (fig. S1, table S8–S11). Matched cohort sizes ranged from 421 to 1,074 per arm (table S6). In this high-risk population, the direction of association was concordant with the LOUE results: sodium channel blockers were associated with a 44% reduction of all-cause dementia (HR = 0.56, 0.39–0.81, Fig. 3A1) and 49% reduction of Alzheimer’s disease (HR = 0.51, 0.33–0.80, Fig. 3A2) compared with SV2A ligands.

**Figure 3.**
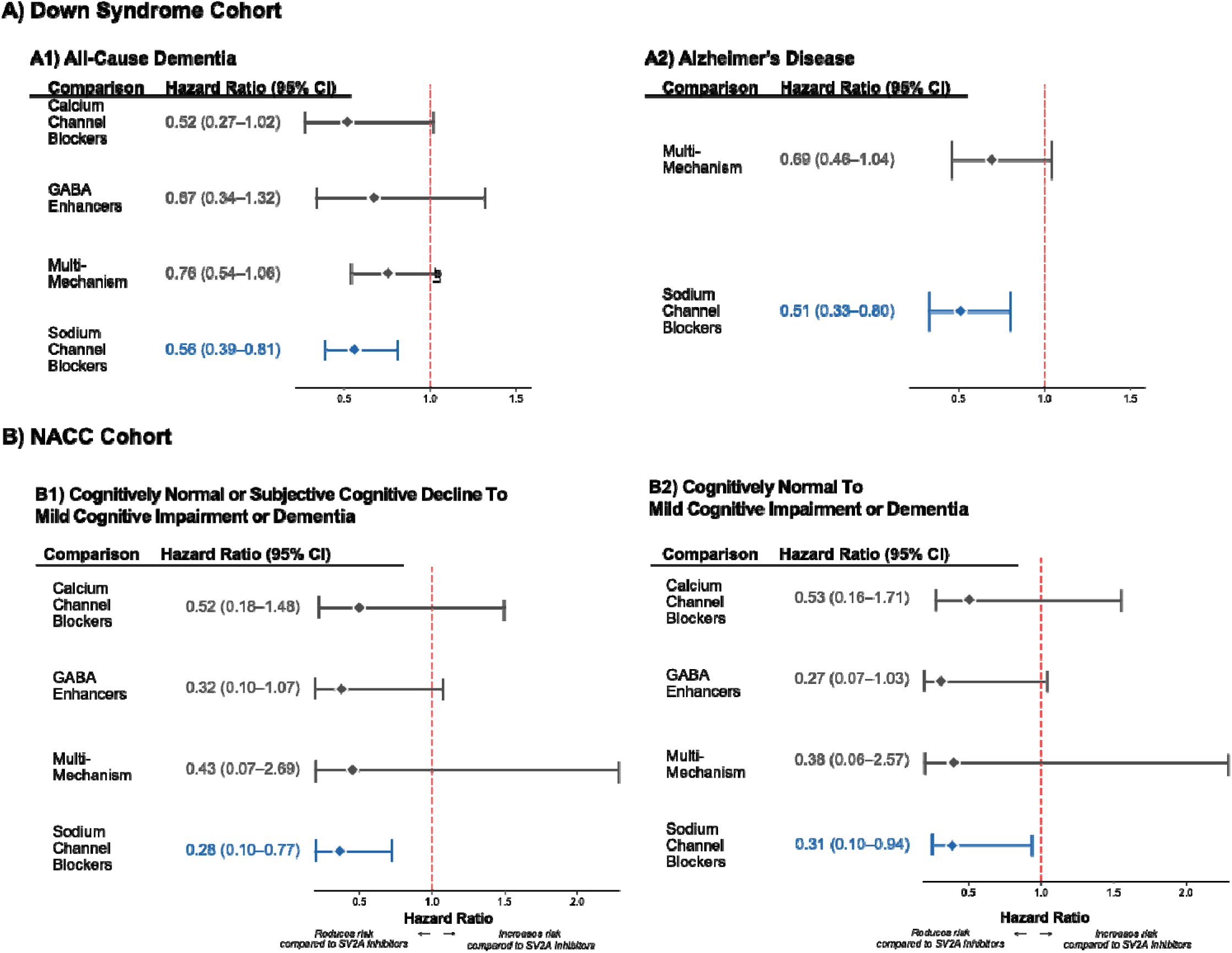
Forest plots of antiseizure medication comparisons in the external validation cohorts. Hazard ratios (95% CI) for dementia and cognitive progression in two external validation cohorts comparing mechanism groups vs SV2A inhibitors. Panels show: **(A) Down Syndrome Cohort**, including (A1) All-cause dementia and (A2) Alzheimer’s disease; and **(B) NACC Cohort**, including progression to MCI or dementia from (B1) Cognitively normal/subjective decline and (B2) Cognitively normal only. Diamond markers indicate hazard ratios; horizontal lines show 95% confidence intervals. Red dashed line at HR=1.0 indicates no difference. Blue indicates a significant protective effect. Gray indicates non-significant results. Per-protocol analysis with follow-up starting at day 366 in Down Syndrome and Intention to Treat analysis in NACC.

In the NACC dataset, 279 individuals with late-onset epilepsy maintained on ASM monotherapy met the eligibility criteria for validation (table S33–S35, fig. S2). Baseline cognitive measures at time zero did not differ significantly across mechanistic groups (Kruskal–Wallis p > 0.15 for all measures; pairwise Mann–Whitney U tests non-significant), confirming comparable cognitive status at study entry (table S36–S37, fig. S3). Using an adjusted intention-to-treat framework anchored to the first eligible visit on monotherapy, sodium channel blockers were associated with a 72% reduction of mild cognitive impairment (MCI) or dementia compared with SV2A ligands (HR = 0.28, 0.10–0.77, Fig. 3B1). This endpoint reflects cognitive progression assessed via standardized neuropsychological testing rather than incident diagnostic codes, and the direction and magnitude of the association support the generalizability of our primary findings.

### Individual ASM associations

Following mechanism-based comparisons, we assessed whether individual ASMs differed from levetiracetam in their association with outcomes. Analyses were restricted to agents with ≥1,000 exposed participants before matching (table S35).

Phenytoin exhibited the strongest protective association, showing a 46% reduction in all-cause dementia (HR = 0.54, 0.39–0.74, Fig. 4A) and a 59% reduction in Alzheimer’s disease (HR = 0.41, 0.25–0.67, Fig. 4B) compared with levetiracetam, corresponding to a 10-year number needed to treat of 33. Post hoc power analyses indicated robust power for these comparisons, with 97.0% power for all-cause dementia (151 observed events; 81 required for 80% power) and 95.6% power for Alzheimer’s disease (67 observed events; 40 required).

**Figure 4.**
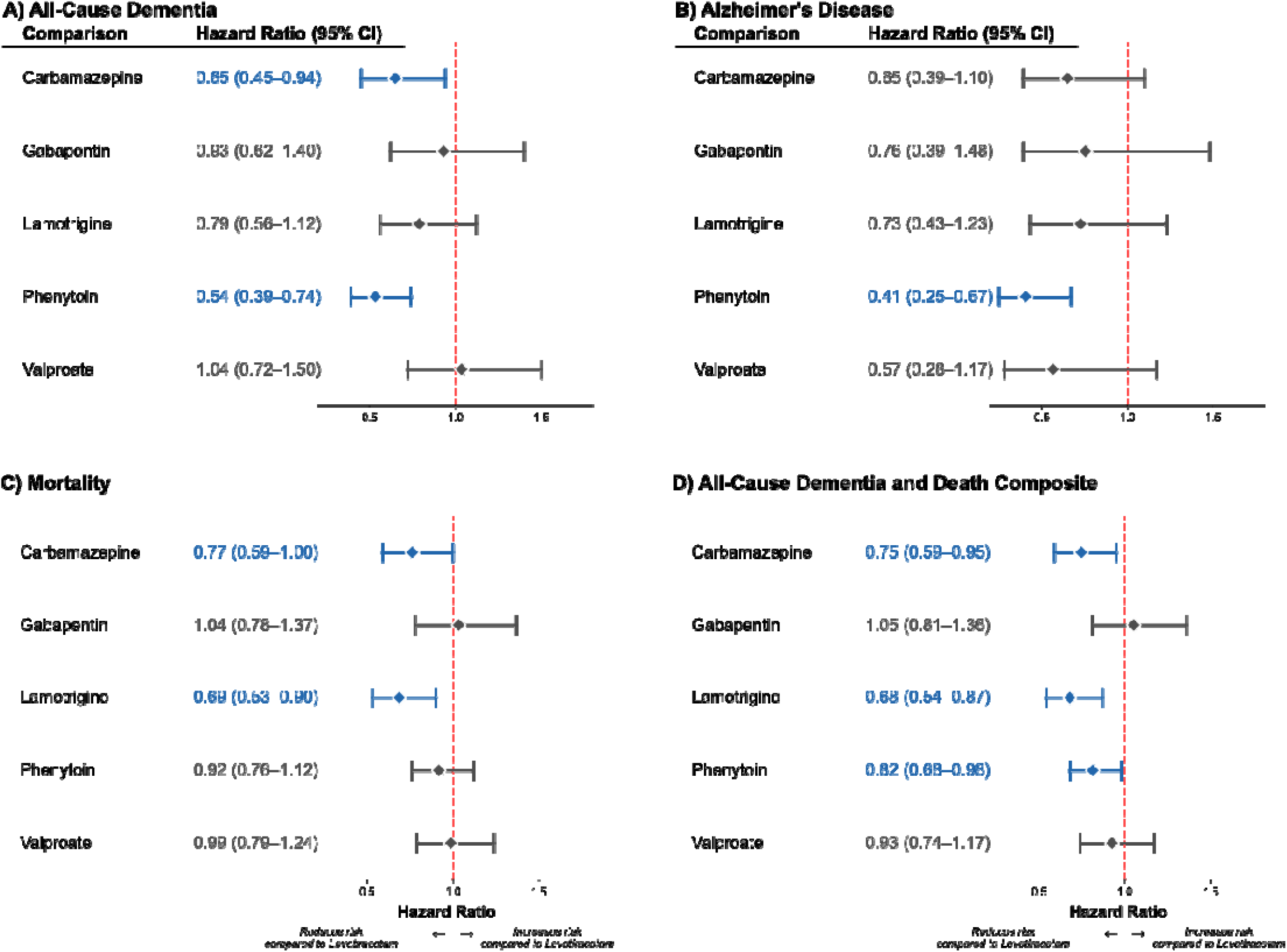
Forest plots of individual antiseizure medications versus levetiracetam. Hazard ratios (95% CI) for dementia and mortality outcomes comparing individual antiseizure medications versus levetiracetam (reference). Panels show: (A) All-cause dementia, (B) Alzheimer’s disease, (C) Mortality, (D) All-cause dementia + death. Diamond markers indicate hazard ratios; horizontal lines show 95% confidence intervals. Red dashed line at HR=1.0 indicates no difference. Blue indicates protective effect, red indicates increased risk, and gray indicates non-significant results. Per-protocol analysis with follow-up starting at day 366. Secondary outcomes (MCI, vascular dementia, glaucoma) shown in supplementary figures.

Carbamazepine was associated with a lower risk of all-cause dementia (HR = 0.65, 0.45–0.94, Fig. 4A) and mortality (HR = 0.77, 0.59–1.00, Fig. 4C), while lamotrigine was associated with lower mortality (HR = 0.69, 0.53–0.90, Fig. 4C). Regarding the composite endpoint of all-cause dementia or mortality, which may reflect the trade-off between efficacy and safety, lamotrigine showed the strongest association (HR = 0.68, 0.54–0.87, Fig. 4D), followed by carbamazepine (HR = 0.75, 0.59–0.95) and phenytoin (HR = 0.82, 0.68–0.98). Gabapentin and valproate showed no significant associations.

In exploratory head-to-head comparisons among the most frequently prescribed sodium channel blockers, phenytoin was associated with a lower risk of Alzheimer’s disease (HR = 0.53, 95% CI 0.31–0.92, Fig. 5B) but with a higher risk of mortality compared with lamotrigine (HR = 1.38, 95% CI 1.12–1.70, Fig. 5C). Additionally, phenytoin was associated with a lower risk of MCI (HR = 0.54, 0.34–0.87, fig. S18A) compared with carbamazepine.

**Figure 5.**
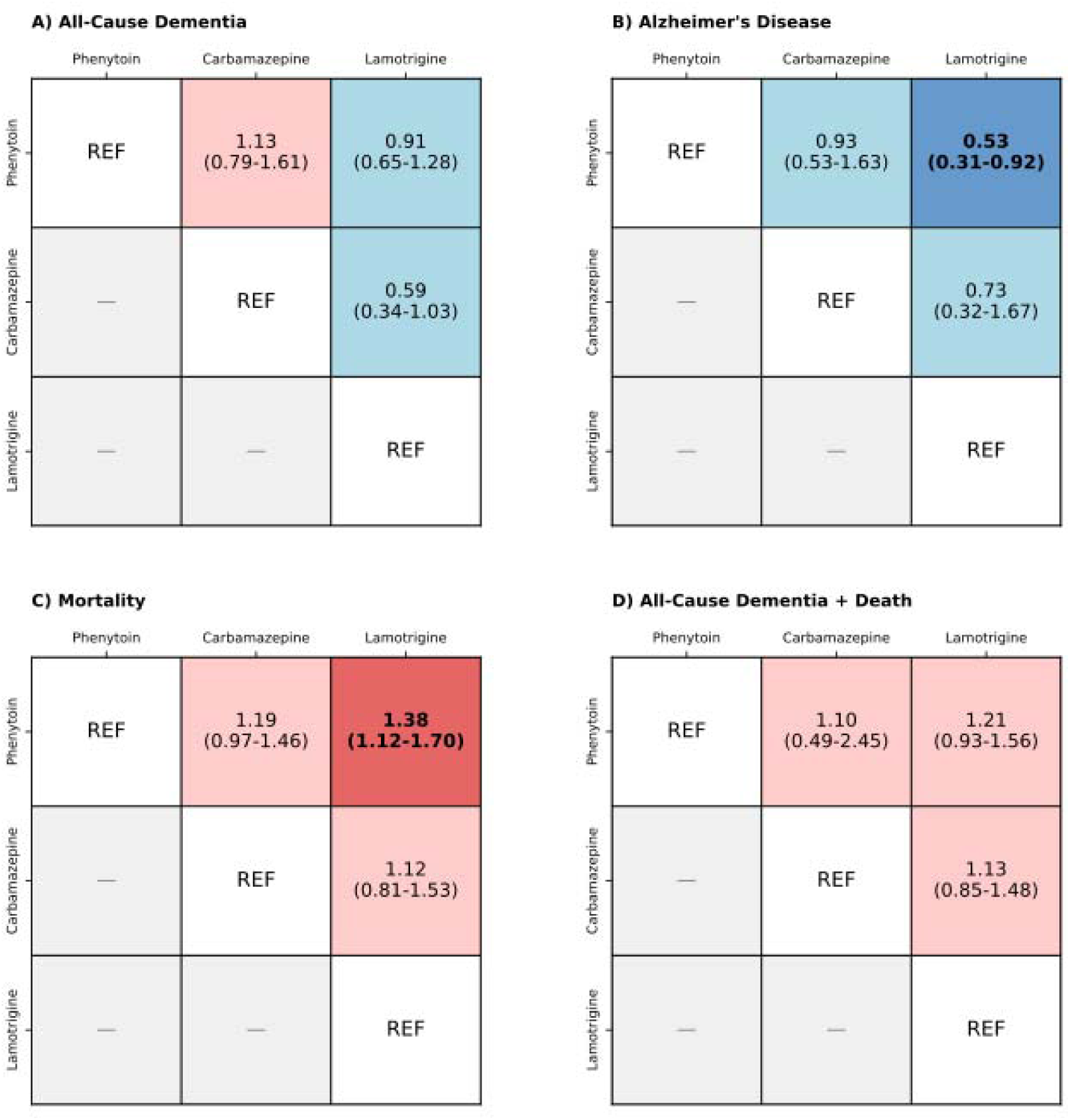
Sodium channel blocker head-to-head comparison matrices. Head-to-head comparison matrices for sodium channel blocker antiseizure medications (Phenytoin, Carbamazepine, Lamotrigine) showing hazard ratios and 95% confidence intervals for primary outcomes. Panels show: (A) All-cause dementia, (B) Alzheimer’s disease, (C) Mortality, (D) All-cause dementia + death. Each cell shows HR (95% CI) where the row drug is compared to the column drug (reference). Cell colors indicate direction of effect: light blue (HR < 1.00, favors row drug), light red (HR > 1.00, favors column drug). Darker shades indicate statistical significance (95% CI excludes 1.00), lighter shades indicate non-significance. Only direct comparisons from source data are shown; inverse comparisons not calculated.

### Secondary and sensitivity analyses

To contextualize these findings, we examined specific dementia phenotypes, earlier cognitive endpoints, and negative-control outcomes designed to probe residual confounding.

Analyzing MCI as an earlier cognitive endpoint, we found that phenytoin (HR = 0.48, 0.26–0.89, fig. S14A) and carbamazepine (HR = 0.49, 0.25–0.95) were associated with reduced risk, whereas the sodium channel blocker class did not show a significant reduction (HR = 0.83, 0.59–1.16, fig. S13A). However, the class was associated with a lower risk of vascular dementia (HR = 0.59, 0.39–0.89, fig. S13B). Neither glaucoma nor frailty markers (negative-control outcomes) showed an association with the relevant ASM strategies, supporting the specificity of the observed dementia associations (fig. S13C–D, fig. S14C–D).

We performed prespecified sensitivity analyses to evaluate robustness against reverse causation, competing risks, and alternative outcome definitions. Extending the burn-in period to 3 years preserved protective associations for sodium channel blockers (all-cause dementia: HR = 0.74, 0.57–0.96; Alzheimer’s disease: HR = 0.67, 0.45–1.00; MCI: HR = 0.65, 0.42–1.00; vascular dementia: HR = 0.51, 0.29–0.89), while negative control outcomes remained non-significant (fig. S15). Results were comparable when shortening the burn-in period to 1 month (fig. S16). Results of survivor-restricted analyses are detailed in the Supplement (fig. S17).

## DISCUSSION

In this large-scale target trial emulation involving adults with late-onset unexplained epilepsy (LOUE), our principal finding is that initial monotherapy with sodium channel blockers was associated with a 27% lower risk of incident all-cause dementia and a 34% reduction in Alzheimer’s disease compared with SV2A ligand therapy. These associations persisted after rigorous adjustment for comorbidities, frailty, and healthcare utilization, and were robust to sensitivity analyses addressing reverse causation and competing risks. Similar protective signals were observed in a Down syndrome cohort and validated externally in the NACC dataset. Direct comparisons among commonly prescribed sodium channel blockers revealed relevant differences: phenytoin was associated with a lower hazard of Alzheimer’s disease compared with lamotrigine, whereas lamotrigine was associated with a survival benefit, highlighting potential efficacy-safety trade-offs in ASM selection (20).

LOUE represents a critical intervention window, often emerging years before clinical dementia. Our findings suggest that ASM selection in this high-risk population is not neutral regarding long-term cognitive trajectories. Our results are in line with the hypothesis that aberrant neural network activity is a driver of neurodegenerative progression rather than merely a symptom. LOUE may mark a “tipping point” in the aging brain, where synaptic hyperexcitability accelerates proteinopathy and cognitive decline (1, 2). Pharmacological dampening of intrinsic neuronal excitability could plausibly slow the transition from concealed network dysfunction to overt dementia (21).

Confidence in this signal is strengthened by the generalization of our primary findings across disparate cohorts and outcomes. The association between sodium channel blockers and reduced dementia risk was replicated in a Down syndrome cohort, a population with a distinct genetic etiology for Alzheimer’s disease and a high baseline risk of early-onset dementia (3).

Furthermore, external validation in the NACC dataset, which utilizes standardized neuropsychological assessments rather than administrative codes, confirmed that sodium channel blockers were associated with a slower progression to mild cognitive impairment or dementia. The consistency of the effect direction across these settings, despite their differing ascertainment methods and confounding structures, reduces the likelihood that the primary finding is solely an artifact of specific selection biases in electronic health records, although residual confounding cannot be entirely excluded.

This study expands upon literature linking network hyperexcitability to cognitive decline. Translational research indicates that hippocampal hyperactivity precedes amyloid plaque deposition and that dampening this activity rescues cognitive deficits in Alzheimer’s models (5, 24). While clinical translation has largely focused on levetiracetam (8, 9, 25), trials have faced recruitment challenges or failed to show clear cognitive benefits in primary outcomes (4, 8, 9). Previous observational studies on ASM risks yielded mixed results, often limited by prevalent-user designs or lack of mechanistic stratification (10, 11). By applying a robust causal inference framework, we provide the first large-scale comparative evidence shifting focus from levetiracetam toward sodium channel blockers as potential cognitive modifiers in LOUE. Mechanistically, the potential benefit of sodium channel blockers may align with their ability to inhibit Nav1.6-mediated high-frequency repetitive firing, consequently breaking the vicious cycle of excitability and amyloid-β release (26). Recent animal studies demonstrate that reducing Nav1.6 activity suppresses intracellular calcium overload and downstream BACE1 transcription, thereby decreasing amyloid-β production independent of plaque burden (27, 28). Interstitial amyloid-β levels and tau propagation are dynamically regulated by synaptic activity, and sodium channel blockade has been shown to profoundly reduce extracellular amyloid concentrations in vivo (12, 13, 15, 29). Beyond direct effects on neuronal excitability, preclinical evidence supports additional neuroprotective mechanisms including autophagy activation (30), mitochondrial stabilization (31), sleep restoration (32), and reduced microglial activation (33). Agent-specific differences in these pleiotropic effects may partly explain the observed hierarchy among sodium channel blockers.

While improved seizure control could theoretically contribute to the observed benefit, several factors argue against this interpretation. First, seizure-related healthcare utilization, a proxy for seizure control, did not differ between treatment groups. Second, LOUE typically presents as focal seizures that are highly responsive to ASM treatment (22), with drug-resistant epilepsy developing in fewer than 10% of cases (23). By restricting our analysis to individuals maintained on monotherapy, we likely selected for a population with uniformly good seizure control, minimizing confounding by disease severity. Alternatively, these findings could reflect non-neuronal mechanisms, such as vascular protection or anti-inflammatory effects. Finally, although residual channeling bias cannot be fully excluded, the robustness of our findings in the 3-year burn-in analysis suggests that simple reverse causation is unlikely to explain the association.

The assessment of individual agents reinforces the notion that the effects on dementia risk are likely attributable to the mechanistic group of sodium channel blockers rather than isolated agents. Nonetheless, the three most widely prescribed sodium channel blockers revealed divergent efficacy and safety profiles. Head-to-head comparisons of these three agents favor phenytoin over lamotrigine for dementia prevention. Phenytoin exhibited particularly robust effects on Alzheimer’s disease. This distinct profile may reflect pleiotropic mechanisms beyond sodium channel modulation; phenytoin inhibits receptor-interacting protein kinase 1 (RIPK1) (16, 18), a key mediator of necroptosis and neuroinflammation, highly activated in Alzheimer’s brains (18). It also inhibits BACE1, the rate-limiting enzyme in amyloid-β production (34), and potentially modulates the TREK-1 potassium channel, recently implicated in amyloid-induced hyperexcitability (35). Lamotrigine also exerts multiple neuroprotective actions, including suppression of network hyperexcitability, reduction of amyloid-β generation (36), attenuation of neuroinflammation (37), preservation of synaptic integrity (36, 38), upregulation of neurotrophic factors such as BDNF and NGF (38), and modulation of hyperpolarization-activated cyclic nucleotide-gated (HCN) channels (30). These insights may highlight specific pathways for therapeutic development.

However, the clinical prioritization of these agents is complicated by safety signals and pharmacological properties in older adults. We observed a significantly increased mortality risk associated with phenytoin compared with lamotrigine, whereas lamotrigine was associated with the most favorable effect size for the composite endpoint of all-cause dementia or death. The use of phenytoin and carbamazepine is further constrained by their strong hepatic enzyme induction, which risks bone health and interferes with common comedications such as statins and direct oral anticoagulants, factors that have driven a shift toward non-enzyme-inducing ASMs (39).

Additionally, phenytoin has a narrow therapeutic index and is associated with gingival hyperplasia, cerebellar atrophy, and cognitive adverse effects. These non-trivial efficacy-safety trade-offs present a clinical dilemma: balancing the potential for disease modification against the imperative for safety and tolerability.

Future prospective studies must therefore rigorously evaluate these competing priorities to determine the optimal therapeutic strategy for LOUE. Furthermore, our results offer a data-driven roadmap for drug repurposing, a rapid and cost-effective strategy to address the urgent global burden of dementia. By systematically comparing mechanistic classes and individual agents, this work provides an evidence-based framework to prioritize specific compounds for translation into pre-clinical Alzheimer’s models and clinical prevention trials.

Our study has limitations. First, the restriction to patients with LOUE without baseline cognitive impairment limits generalizability to individuals with pre-existing dementia or those without epilepsy. Second, despite robust propensity-score matching, residual unmeasured confounding cannot be ruled out; however, the consistency of results across negative control outcomes (glaucoma, frailty, healthcare utilization) and disparate validation cohorts (Down syndrome, NACC) argues against confounding as the sole explanation of our findings. Third, residual confounding by epilepsy severity, seizure frequency, and baseline cognitive status may persist, as standardized metrics of seizure frequency, electroencephalography, and detailed neuropsychological performance are not uniformly captured in federated electronic health records. However, our restriction to patients maintained on ASM monotherapy likely reduces the confounding influence of disease burden. Furthermore, in the NACC validation cohort, baseline cognitive metrics did not differ across mechanistic groups, suggesting that between-group differences in cognitive outcomes are unlikely to be driven by baseline cognitive imbalances.

Fourth, relying on administrative diagnostic codes introduces potential misclassification and variability in diagnostic latency, although validation using the NACC dataset, with standardized neuropsychological assessments, helps mitigate this concern. Fifth, the lack of findings in smaller ASM classes may reflect limited power. Finally, while our per-protocol design allows for the estimation of biological effects under sustained ASM monotherapy, the findings may not fully generalize to real-world clinical scenarios in which treatment switching or polytherapy are permissible.

In conclusion, this target trial emulation study suggests that initial ASM selection in late-onset unexplained epilepsy is associated with differential dementia risk. The consistent benefit of sodium channel blockers across diverse cohorts supports the hypothesis that modulating neuronal excitability with these agents may modify the neurodegenerative trajectory. While confirmatory randomized trials are needed to establish causality, these findings hold immediate relevance for clinical decision-making. Clinicians might consider these associations when counselling patients with LOUE who are concerned about their long-term cognitive trajectory, particularly when choosing between equipotent agents.

## MATERIALS AND METHODS

### Experimental Design

We conducted a retrospective cohort study using a new-user, active-comparator design emulating a parallel-group target trial to evaluate whether selection of antiseizure medication (ASM) monotherapy modifies dementia risk in adults with late-onset epilepsy of unexplained etiology (LOUE). The analytic framework followed target-trial emulation principles and was reported in accordance with STROBE and the TARGET recommendations (40).

Data were obtained from the TriNetX Global Collaborative Network (https://trinetx.com/) (41), a federated platform of de-identified real-world international health data harmonized across participating healthcare organizations. At the time of the search (November 2025), the network included 157 organizations across multiple world regions and contained >250 million individuals with longitudinal, date-stamped observations. TriNetX captures structured demographics, diagnoses (ICD-10-CM), medications (RxNorm), procedures, laboratory values, and mortality. Data refresh occurs continuously, and de-identification complies with HIPAA Privacy Rule (§164.514[b][1]).

### Target Trial Specification

We applied a target trial emulation framework to estimate the effect of ASM selection on dementia risk. We specified the protocol of the target trial, which enrolled adults with incident LOUE initiating monotherapy with no prior dementia-spectrum diagnoses or anti-dementia medications, and no competing neurologic or systemic conditions strongly associated with secondary epilepsy or dementia. Eligible individuals were assigned to an SV2A-ligand strategy or a comparator ASM strategy. Time zero was defined as the date when eligibility criteria were met (incident epilepsy diagnosis and ASM monotherapy initiation). We emulated this trial in the TriNetX real-world international health dataset by strictly aligning eligibility, exposure definition, and follow-up with this protocol, and used propensity-score matching to address confounding inherent to non-randomized treatment assignment.

### Study Population

We identified individuals with epilepsy diagnoses (ICD-10-CM G40.*) first recorded at age ≥55 years and required at least two epilepsy diagnosis instances to improve the accuracy of LOUE ascertainment. This validated case ascertainment strategy, when paired with ASM prescription, achieves 80–90% positive predictive value (PPV) and sensitivity estimates for capturing epilepsy within health data, and negative predictive values (NPV) and specificity estimates approach 100% (42, 43). To operationalize “unexplained” late-onset epilepsy and mitigate reverse causation, we excluded individuals with acute symptomatic seizures, prior ASM exposure, dementia (Alzheimer’s disease, vascular dementia, Lewy body dementia, frontotemporal dementia), mild cognitive impairment (MCI), or anti-dementia medication use (e.g., donepezil, rivastigmine, galantamine, memantine, lecanemab, aducanumab, donanemab) before index or during the prespecified burn-in period.

We further excluded conditions associated with secondary epilepsy in older adults or independently associated with dementia risk, including acquired structural brain lesions; central nervous system infections; postconcussion syndrome/traumatic brain injury; hydrocephalus; arteriovenous or cavernous malformations; ischemic or hemorrhagic stroke; sequelae of inflammatory CNS disease; other neurodegenerative conditions; end-stage systemic comorbidities (end-stage renal disease, end-stage heart failure, chronic hepatic failure); and severe psychiatric or substance-use disorders before index or during the burn-in period. Full inclusion and exclusion code lists are provided in table S2–S3.

### Exposure Groups

We compared SV2A-ligand monotherapy (reference strategy: levetiracetam and brivaracetam) versus alternative ASM monotherapies grouped by primary mechanism of action: (i) sodium channel blockers: carbamazepine, oxcarbazepine, eslicarbazepine, lamotrigine, lacosamide, phenytoin; (ii) calcium channel blockers: gabapentin, pregabalin; (iii) GABAergic enhancers: phenobarbital, clobazam, clonazepam; and (iv) multiple mechanisms: valproic acid, topiramate, zonisamide.

SV2A ligands were prespecified as the reference because levetiracetam is commonly prescribed in LOUE, maximizing statistical power for active-comparator contrasts. Medications were identified using RxNorm codes. To better capture chronic outpatient exposure and reduce confounding by acute-care indications, we restricted exposure to oral formulations when agents were available in both oral and intravenous forms. New-user monotherapy was defined as receipt of at least two prescriptions of the assigned ASM within a 24-month period, with no evidence of current or prior exposure to any other ASM (44).

### Time Zero, Burn-In Period, and Follow-up

Time zero (index date) was defined as the first day on which an individual met both: (i) at least two recorded epilepsy diagnoses, and (ii) at least two prescriptions for the assigned ASM, consistent with the new-user monotherapy definition. To reduce reverse causation, we prespecified a 365-day burn-in period after time zero during which outcomes were not counted. Follow-up began on day 366 and continued until the first occurrence of an outcome event, death, loss of electronic health record activity (defined as absence of any recorded encounter/diagnosis/medication/procedure for ≥90 consecutive days before data extraction), or administrative end of data availability (September 2025).

### Outcomes

Primary outcomes were: (i) all-cause dementia, defined as the first recorded diagnosis of Alzheimer’s disease, Lewy body dementia, frontotemporal dementia, vascular dementia, unspecified dementia, or dementia in other diseases classified elsewhere; (ii) Alzheimer’s disease; (iii) all-cause mortality; and (iv) a combined endpoint of all-cause dementia or mortality. Secondary outcomes were vascular dementia and MCI. We evaluated two negative control outcomes intended to probe residual confounding: (i) incident glaucoma and (ii) frailty markers. Outcome codes can be found in table S4.

### Covariates and Propensity-Score Matching

To mitigate confounding, we adjusted for established determinants of dementia risk, treatment selection, clinical complexity, frailty, and healthcare utilization. Baseline covariates were ascertained at any time before the index date and included age at index, sex, race/ethnicity, vascular risk factors (hypertension, diabetes, hyperlipidemia, sleep disorders), sensory impairment (vision/hearing problems), and concomitant medication classes including antidepressants, antipsychotics, benzodiazepines, cardiovascular agents, statins, and anticoagulants or antiplatelets. We additionally incorporated frailty metrics and healthcare utilization indices (45, 46). Full covariate lists are provided in table S5. Propensity scores were estimated using logistic regression (47, 48). Matching was performed on a 1:1 basis using greedy nearest-neighbor matching without replacement, with a caliper width of 0.1 pooled standard deviations (49, 50). Each comparator group was independently matched to the reference cohort, allowing SV2A initiators to serve as comparators across multiple contrasts.

### Statistical Analysis

In matched cohorts, hazard ratios (HRs) and 95% confidence intervals (CIs) were estimated using Cox proportional hazards regression. SV2A ligands served as the reference; thus, an HR >1 indicated a higher risk of dementia relative to the reference. Results were summarized in forest plots. For comparisons demonstrating statistical significance, absolute treatment effects were quantified by calculating the number needed to treat (NNT) at the 10-year follow-up, defined as the reciprocal of the absolute risk difference derived from Kaplan–Meier survival estimates. Post hoc power analyses were performed using the Schoenfeld formula. An SMD threshold of ≤0.1 was used to indicate when covariates were well matched (19). All cohort construction, matching, and time-to-event analyses were performed within the TriNetX Analytics environment (version October 2025). Outputs were exported for downstream formatting and visualization using Python (version 3.11.9).

### Secondary and Sensitivity Analyses

We conducted prespecified sensitivity analyses to assess robustness under alternative assumptions, including: (i) a shorter burn-in period (1 month); (ii) an extended burn-in period (1 to 3 years) to further mitigate reverse causation; (iii) competing risk of death assessed by repeating analyses after excluding individuals who died during follow-up (survivor-restricted analysis); and (iv) MCI as an earlier cognitive outcome using the same matching and survival framework.

### Validation Analyses

Down syndrome TriNetX cohort: To evaluate generalizability in a population with high baseline dementia risk, we identified individuals with Down syndrome (ICD-10 Q90.*) and repeated the exposure definitions and analytic workflow. No comorbidity-based exclusions were applied to maximize sample size. Inclusion and exclusion criteria codes can be found in table S9–S11.

NACC cohort: The NACC maintains a multicenter longitudinal database of participants enrolled across 34 National Institute on Aging–funded Alzheimer’s Disease Research Centers with standardized clinical assessments, neuropsychological testing, and neuroimaging, providing an independent, well-characterized cohort for external validation. We included data for UDS visits conducted between September 2005 and September 2024. All participants provided informed consent at their respective ADRC sites. Epilepsy status was defined using a composite criterion of either documented epilepsy diagnosis or recorded seizure history together with concurrent ASM use at the same visit. Cognitive status was derived from NACCUDSD codes and categorized as cognitively normal, subjective cognitive decline, MCI, or dementia, with time zero defined as the first visit with ASM exposure at which cognition was normal/subjective cognitive decline. The primary outcome was progression to MCI or dementia at a subsequent visit. Cox proportional hazards regression compared cognitive progression risk between SV2A ligands and other ASMs grouped by mechanism. Propensity score matching was not performed due to limited sample size, but effect estimates were adjusted for epilepsy status, sex, years of education, and age at inclusion.

## LIST OF SUPPLEMENTARY MATERIALS

### Materials and Methods

Fig. S1. Patient flow diagram for Down syndrome cohort

Fig. S2. Patient flow diagram for NACC cohort

Fig. S3. Baseline cognitive measures across ASM groups in the NACC cohort

Fig. S4–S12. Covariate balance plots for mechanism-group comparisons (standardized mean differences)

Fig. S13. Secondary outcomes for mechanism group comparisons (MCI, vascular dementia, glaucoma, frailty)

Fig. S14. Secondary outcomes for individual ASM comparisons vs levetiracetam

Fig. S15. Sensitivity analysis: 3-year burn-in period

Fig. S16. Sensitivity analysis: 1-month burn-in period

Fig. S17. Survivor-restricted analyses

Fig. S18. Head-to-head comparisons of sodium channel blockers: secondary outcomes

Table S1. Target Trial Protocol and Emulation Using Electronic Health Records

Table S2. Inclusion Criteria

Table S3. Exclusion Criteria

Table S4. Study Outcomes

Table S5. Covariates for Propensity Score Matching

Table S6. Sample Sizes Before and After Propensity Score Matching

Table S7. Mean Follow-up Time by Comparison Group

Table S8. Baseline Characteristics of Down Syndrome Cohort

Table S9. Target Trial Protocol and Emulation for Down Syndrome Cohort

Table S10. Inclusion Criteria for Down Syndrome Cohort

Table S11. Covariates for Propensity Score Matching in Down Syndrome Cohort

Tables S12–S25. Baseline Characteristics After Matching (mechanism group and individual ASM comparisons, main and 1-month burn-in analyses)

Tables S26–S32. Baseline Characteristics After Matching in Down Syndrome Cohort

Tables S33–S35. NACC cohort characteristics and eligibility

Tables S36–S37. Baseline cognitive measures in NACC cohort

*References (40–50) are cited only in Materials and Methods and Supplementary Materials*.

## Data Availability

https://live.trinetx.com/

## ACKNOWLEDGMENTS

The NACC database is funded by NIA/NIH Grant U24 AG072122. NACC data are contributed by the NIA-funded ADRCs: P30 AG062429 (PI James Brewer, MD, PhD), P30 AG066468 (PI Oscar Lopez, MD), P30 AG062421 (PI Bradley Hyman, MD, PhD), P30 AG066509 (PI Thomas Grabowski, MD), P30 AG066514 (PI Mary Sano, PhD), P30 AG066530 (PI Helena Chui, MD), P30 AG066507 (PI Marilyn Albert, PhD), P30 AG066444 (PI John Morris, MD), P30 AG066518 (PI Jeffrey Kaye, MD), P30 AG066512 (PI Thomas Wisniewski, MD), P30 AG066462 (PI Scott Small, MD), P30 AG072979 (PI David Wolk, MD), P30 AG072972 (PI Charles DeCarli, MD), P30 AG072976 (PI Andrew Saykin, PsyD), P30 AG072975 (PI David Bennett, MD), P30 AG072978 (PI Neil Kowall, MD), P30 AG072977 (PI Robert Vassar, PhD), P30 AG066519 (PI Frank LaFerla, PhD), P30 AG062677 (PI Ronald Petersen, MD, PhD), P30 AG079280 (PI Eric Reiman, MD), P30 AG062422 (PI Gil Rabinovici, MD), P30 AG066511 (PI Allan Levey, MD, PhD), P30 AG072946 (PI Linda Van Eldik, PhD), P30 AG062715 (PI Sanjay Asthana, MD, FRCP), P30 AG072973 (PI Russell Swerdlow, MD), P30 AG066506 (PI Todd Golde, MD, PhD), P30 AG066508 (PI Stephen Strittmatter, MD, PhD), P30 AG066515 (PI Victor Henderson, MD, MS), P30 AG072947 (PI Suzanne Craft, PhD), P30 AG072931 (PI Henry Paulson, MD, PhD), P30 AG066546 (PI Sudha Seshadri, MD), P20 AG068024 (PI Erik Roberson, MD, PhD), P20 AG068053 (PI Justin Miller, PhD), P20 AG068077 (PI Gary Rosenberg, MD), P20 AG068082 (PI Angela Jefferson, PhD), P30 AG072958 (PI Heather Whitson, MD), P30 AG072959 (PI James Leverenz, MD).

## Author contributions

C.F.-A. and M.G. conceptualized the study. C.F.-A. performed the data analysis and wrote the original manuscript draft. T.M., G.Y.H.L., and G.K.M. provided access to the TriNetX data and curated the research resources. C.F.-A., K.M.S., X.Y.T., D.N., D.S., M.W., S.S., H.H.J., T.H., J.Z., T.M., G.Y.H.L., G.K.M., and M.G. provided critical intellectual input and performed a formal review and editing of the manuscript. All authors reviewed and approved the final version of the manuscript.

## Data and materials availability

The clinical and neuropathological data were provided by NACC. These data are available to researchers upon request and approval of a data use agreement via the NACC website (https://naccdata.org). The TriNetX electronic health record data are available to researchers who have a valid license or agreement with TriNetX (https://trinetx.com). All summary data, cohort definitions (including ICD-10 and RxNorm codes), and statistical outputs required to interpret the study findings are provided in the Supplementary Information and Source Data files. Custom code used for external validation is available from the corresponding author upon reasonable request.

**Extended Data Table 1.**
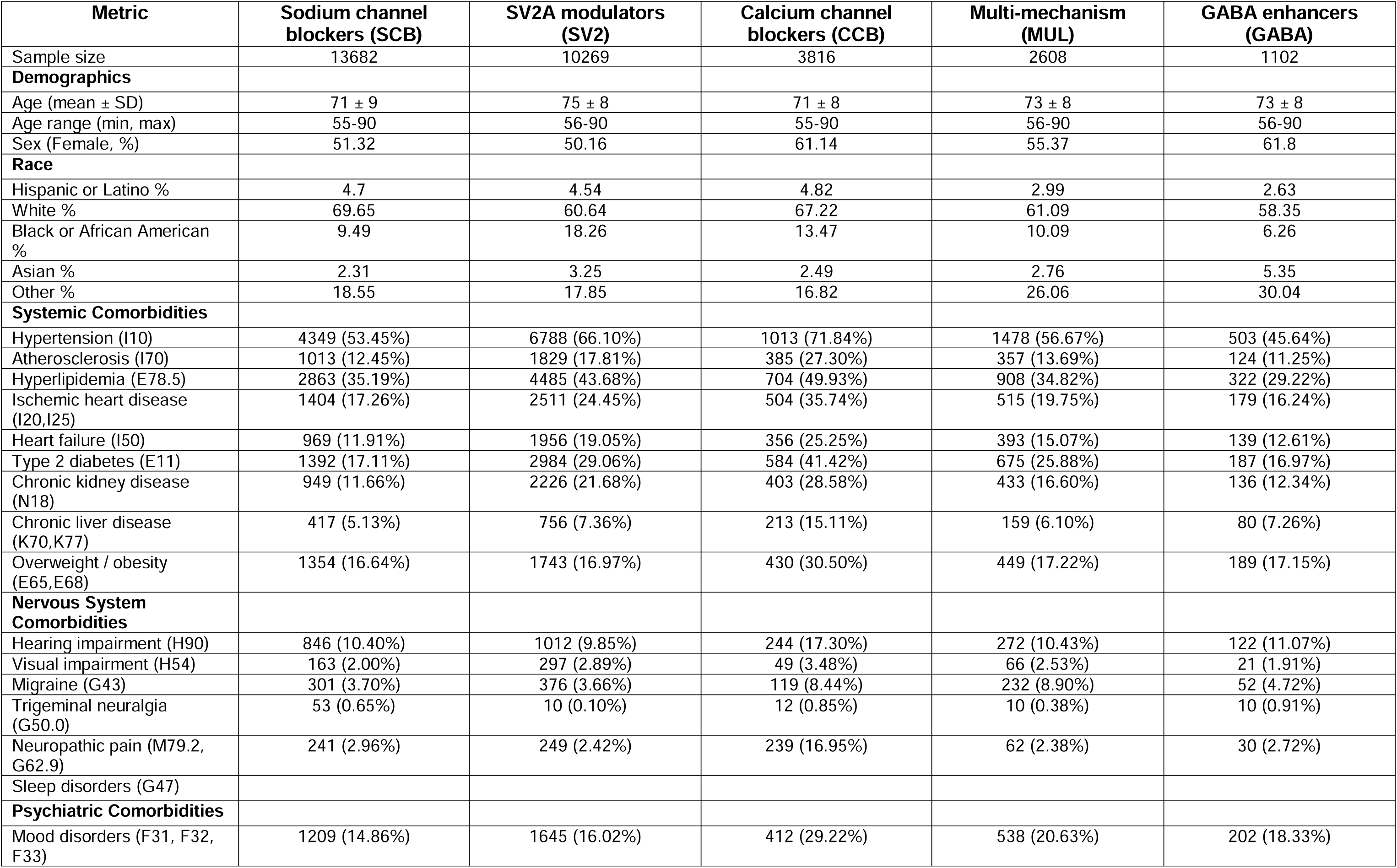
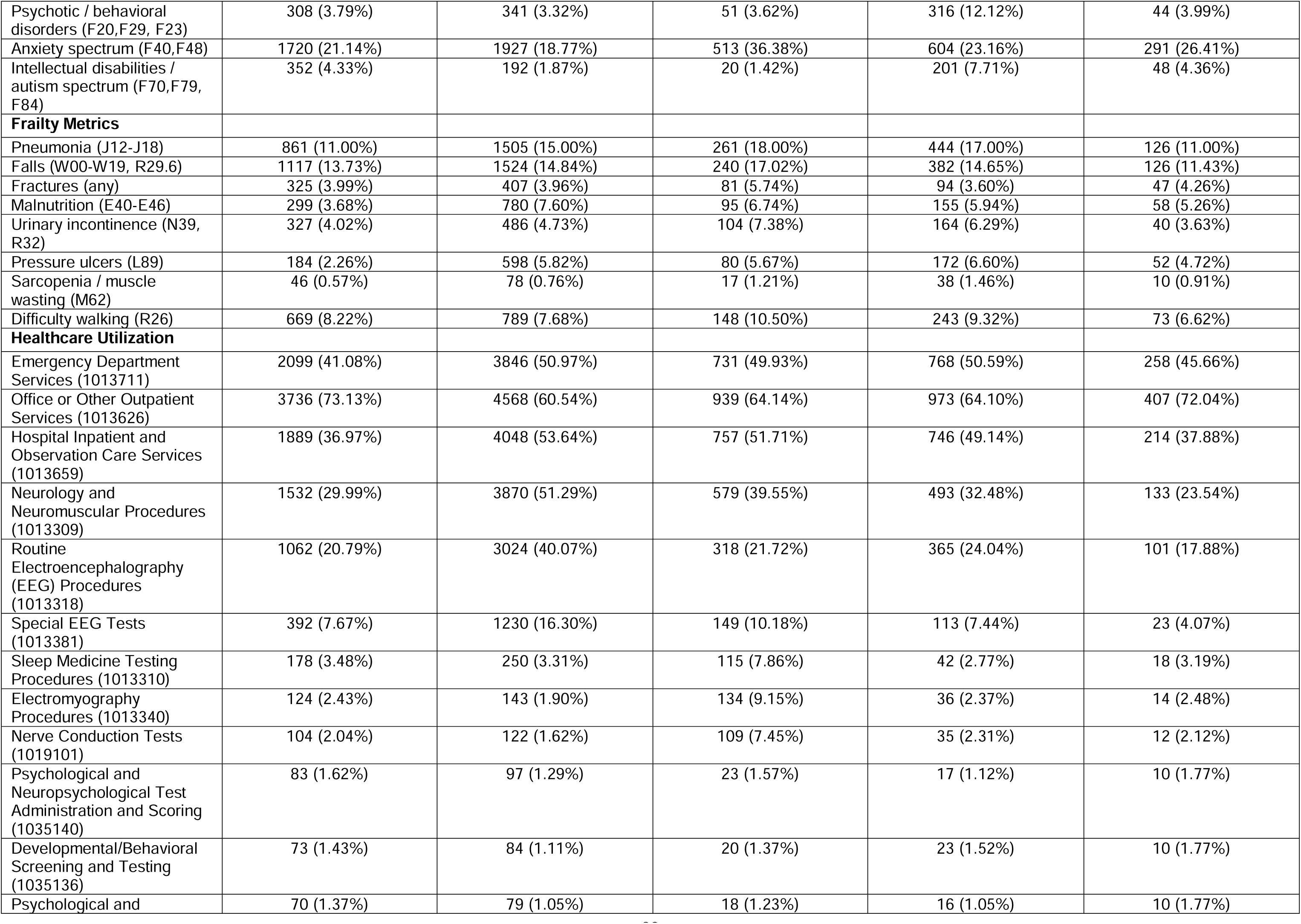
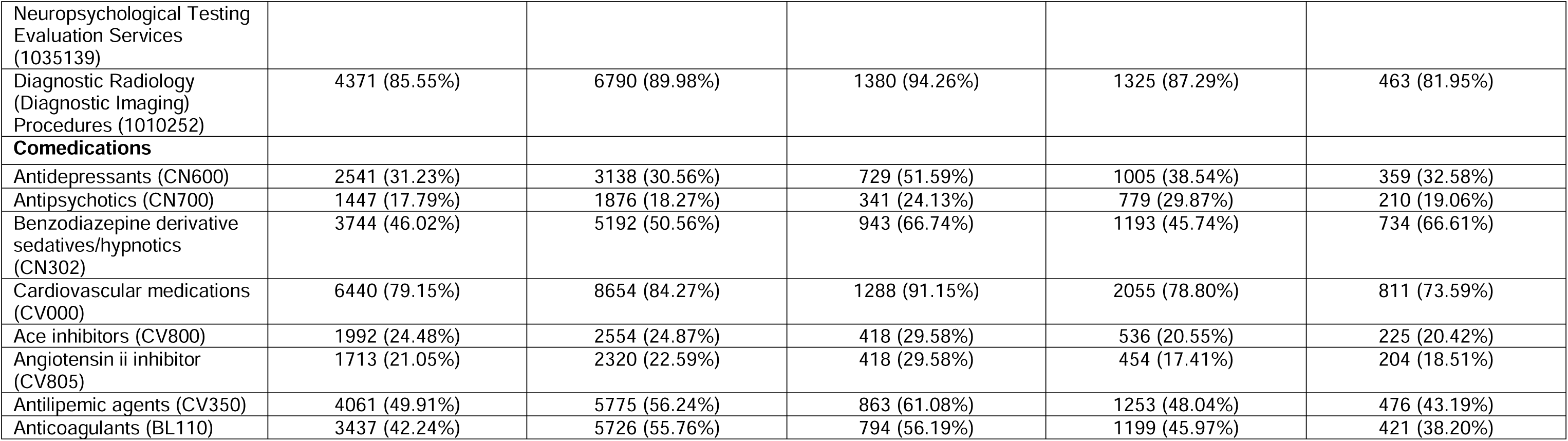
Baseline Characteristics by Mechanism Group.

**Extended Data Table 2.**
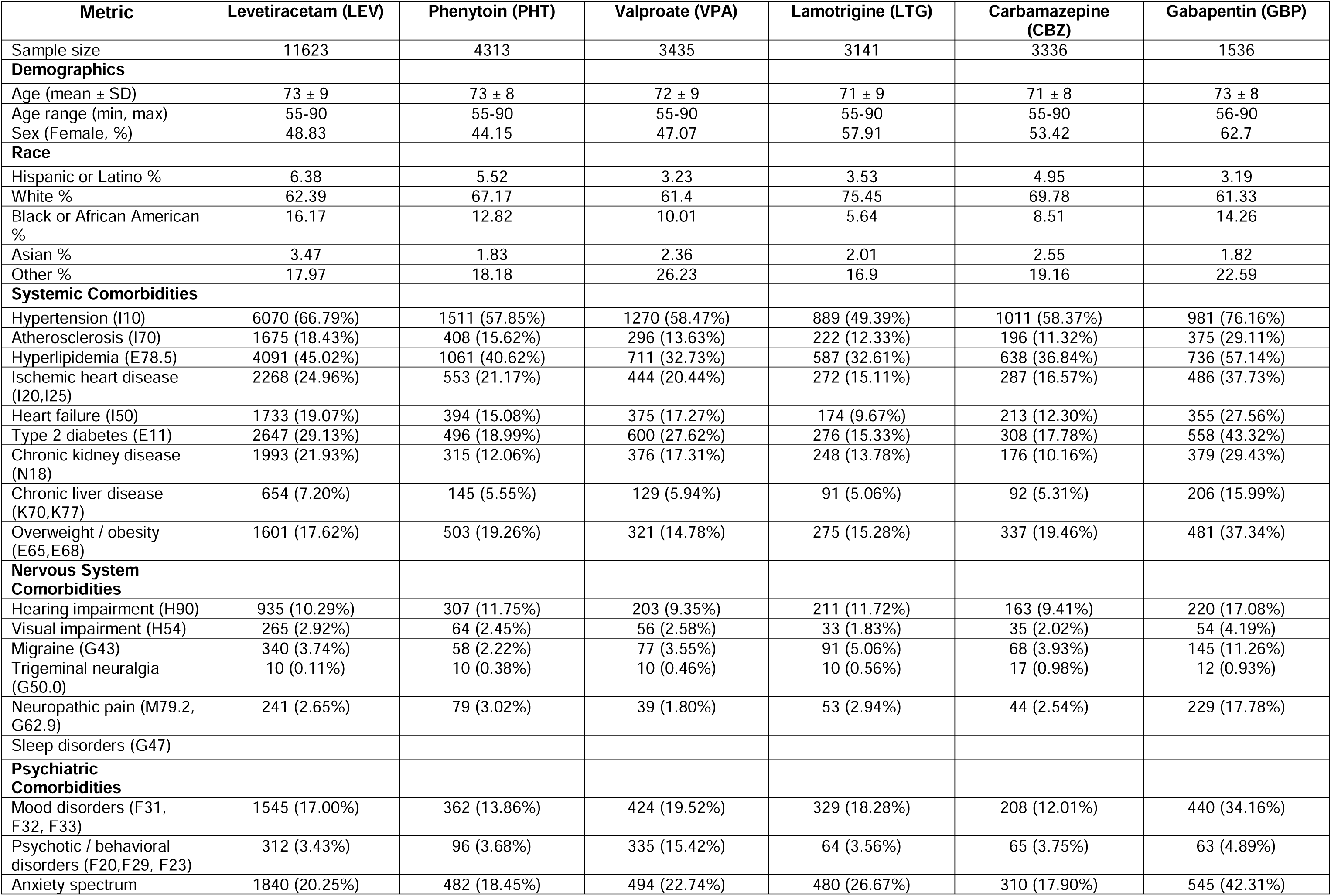
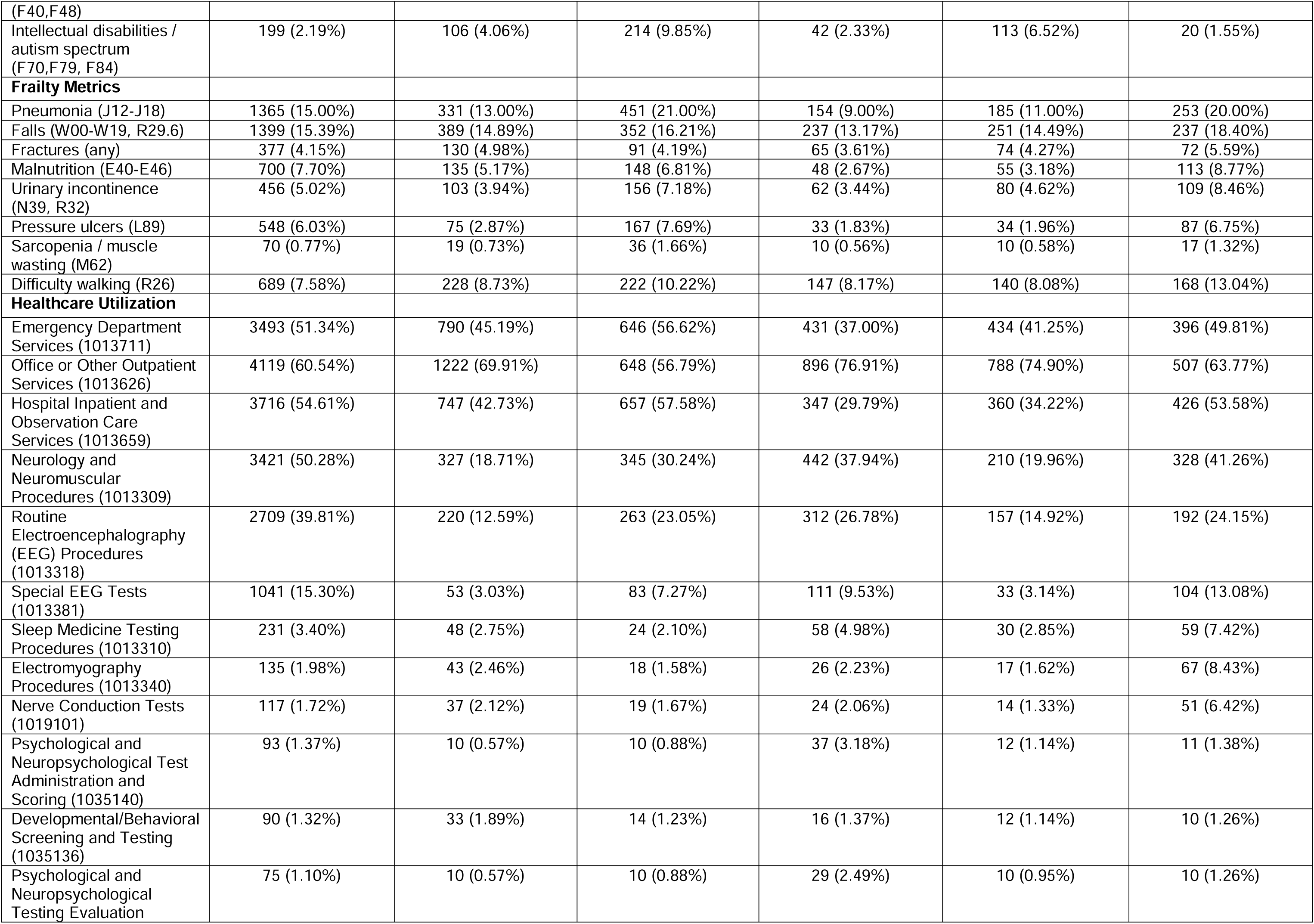
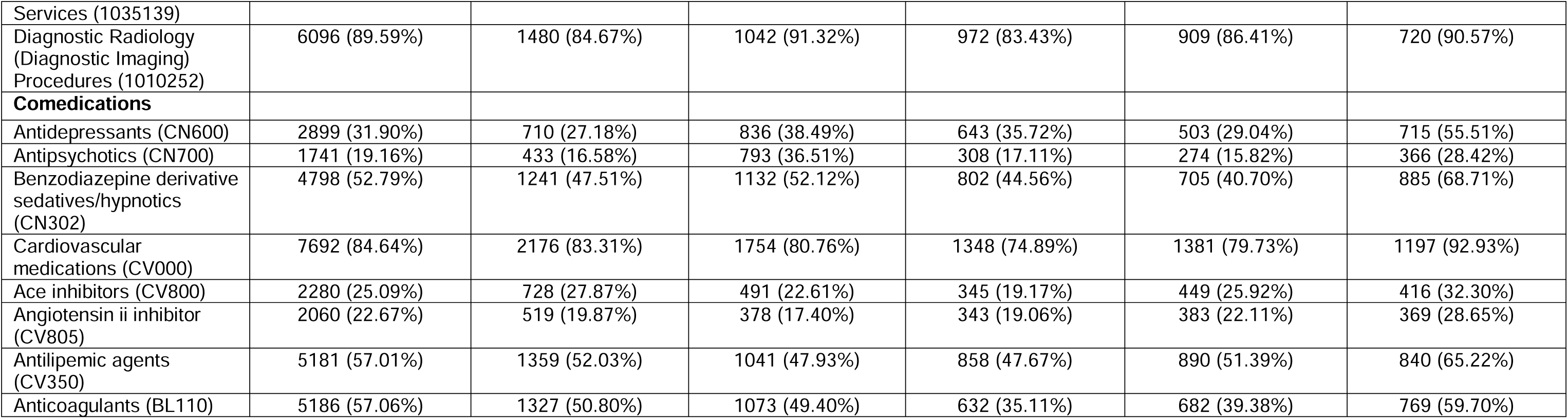
Baseline Characteristics by Individual Antiseizure Medication.

